# County-Year Informatics Model for Annual and Cumulative-Unique Lung Cancer Screening Eligibility in Maryland, 2026–2045

**DOI:** 10.64898/2026.06.15.26355716

**Authors:** Clement Adebamowo, Sally N. Adebamowo

## Abstract

**Purpose:** Population-level lung cancer screening programs require denominators that reflect age, smoking history, geography, and changing eligibility over time. We estimated annual prevalent and 20-year cumulative unique low-dose computed tomography screening eligibility for Maryland residents under alternative screening criteria.

**Methods:** We built a deterministic cohort-cell stock-flow simulation using Maryland county-equivalent jurisdiction projections by age, sex, and race/ethnicity, with ACS socioeconomic/nativity covariates and smoking-history priors for ever-smoked status, pack-years, and quit-years. Scenarios included USPSTF 2013 legacy, USPSTF 2021, ACS 2023/2024, a risk-model-expanded sensitivity, and ever-smoked-only capacity stress tests. Cumulative unique eligibility counted people once at first eligibility rather than summing annual prevalent person-years.

**Results:** Under USPSTF 2021, an estimated 238,346 Maryland residents were eligible in 2026 and 245,326 in 2045. The 20-year cumulative unique denominator was 768,668, whereas naively summing annual prevalent counts produced 4,850,735 person-years, a 6.31-fold overcount. ACS 2023/2024 expanded annual eligibility to 314,616 in 2026 and cumulative unique eligibility to 902,796 by adding remote former smokers. Ever-smoked-only adult eligibility was 1,957,699 in 2026 and 3,383,683 cumulative unique over 20 years.

**Conclusion:** A Maryland statewide screening initiative should plan from cumulative unique eligibility and county-equivalent jurisdiction-specific burden rather than annual prevalence alone. Explicit pack-year and quit-year modeling materially changes statewide and county allocation compared with current-smoking proxy models.

**Context Summary:**

- Key objective: Estimate annual and cumulative unique lung cancer screening eligibility across Maryland county-equivalent jurisdictions under multiple criteria.
- Knowledge generated: The USPSTF 2021 20-year cumulative unique denominator is 768,668 persons, while annual prevalent person-year summation would overcount unique eligible individuals by 6.31-fold.
- Relevance: The model produces reproducible county-year, subgroup, and smoking-history outputs that can support screening capacity planning and equity monitoring.

## Introduction

Lung cancer is the second most common cancer and the leading cause of cancer death in the United States, with an estimated 234,580 new cases and 125,070 deaths projected in 2024.^1-6^ Screening with low-dose computed tomography (LDCT) reduces lung cancer mortality among high-risk individuals.^3,5,7^ Two landmark randomized trials - the National Lung Screening Trial (NLST), which demonstrated a 20% reduction in lung cancer mortality,^7^ and the Dutch-Belgian NELSON trial, which reported a 24% reduction among men^3^ - established the evidentiary basis for population-based LDCT screening programs.^8^

The US Preventive Services Task Force (USPSTF) first recommended annual LDCT screening in 2013 for adults aged 55–80 years with at least 30 pack-years of smoking history,^9^ and in 2021 broadened eligibility to adults aged 50–80 years with at least 20 pack-years.^10^ The American College of Chest Physicians has endorsed similar criteria.^11^ Despite this evidence and guideline endorsement, implementation remains constrained by incomplete risk ascertainment, low uptake,^12^ racial and ethnic disparities in screening adherence,^13-15^ and geographic access barriers.^16,17^

The prevalence of cigarette smoking among US adults has declined to approximately 14% nationally,^18^ yet many former smokers retain substantial cumulative pack-year exposures that qualify them for screening.^19^ Statewide screening programs need population denominators that are both clinically criterion-specific and operationally interpretable. A single annual prevalent count is insufficient for long-horizon planning because people enter, remain in, and exit eligibility over multiple years.

Maryland is heterogeneous in age structure, racial and ethnic composition, smoking prevalence, rurality, and access to imaging capacity.^20^ Current-smoking proxy models can provide rapid approximations, but they are not equivalent to USPSTF eligibility because many qualifying individuals are former smokers with sufficient cumulative pack-years and recent quit dates.^19^ Conversely, the American Cancer Society (ACS) 2023/2024 criteria remove the quit-year exclusion and therefore add remote former smokers who are excluded under USPSTF 2021.^21^

We developed a population-informatics modeling package to estimate annual prevalent and cumulative unique lung cancer screening eligibility in Maryland from 2026 through 2045. The analysis integrates Maryland projection data, county-year estimates, alternative screening criteria, pack-year and quit-year distributions, and subgroup outputs by age, sex, race/ethnicity, county-equivalent jurisdiction, education, income, and nativity.

## Methods

### Population projection spine

The projection spine was organized as annual Maryland resident counts for 2026–2045 across 24 county-equivalent jurisdictions, age group, sex, and race/ethnic origin.^22^ Baltimore City was treated as a county-equivalent jurisdiction. Socioeconomic variables not included in the population projection spine were attached as marginal county-equivalent ACS covariates for subgroup allocation and equity monitoring (Table 1).^23^

**Table 1.**
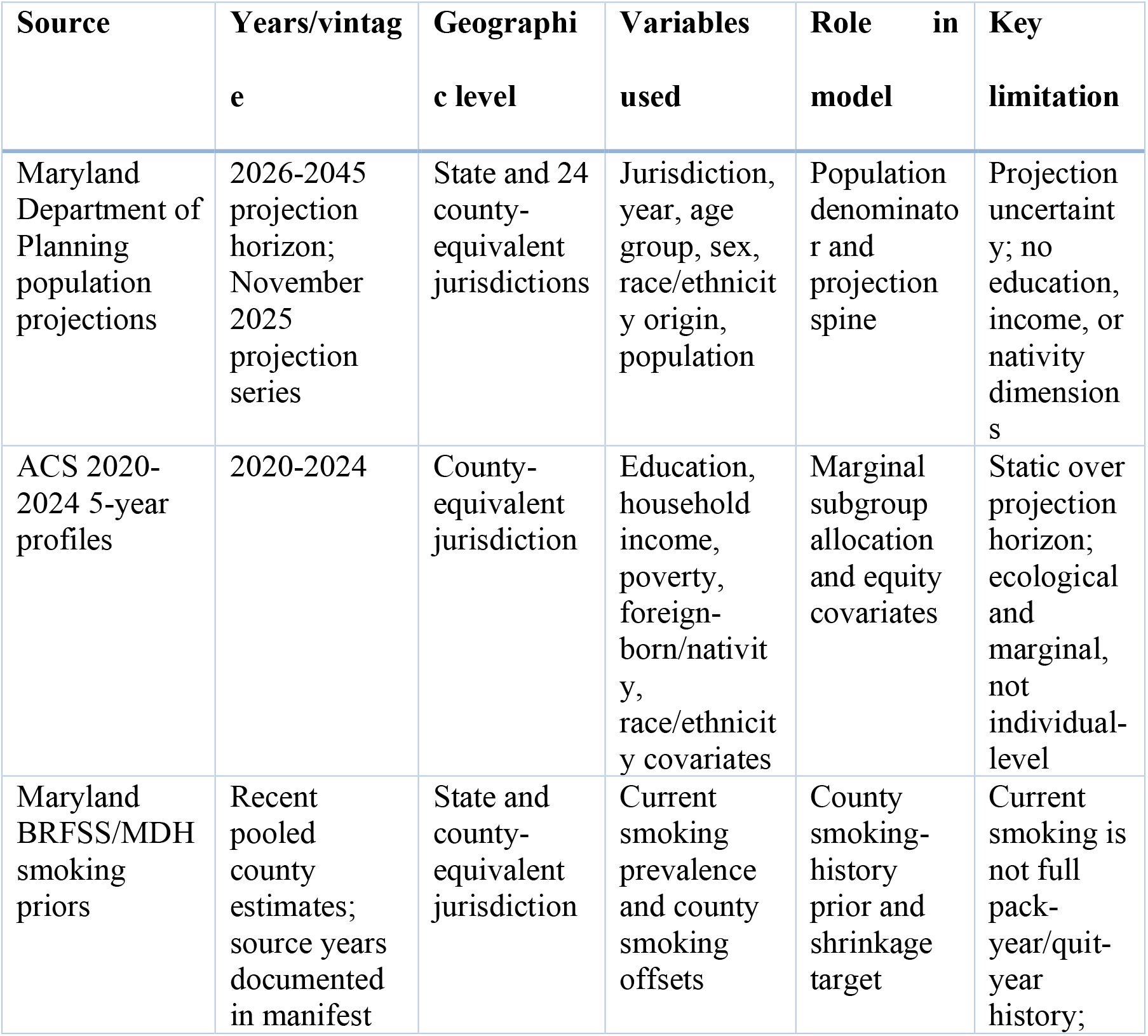
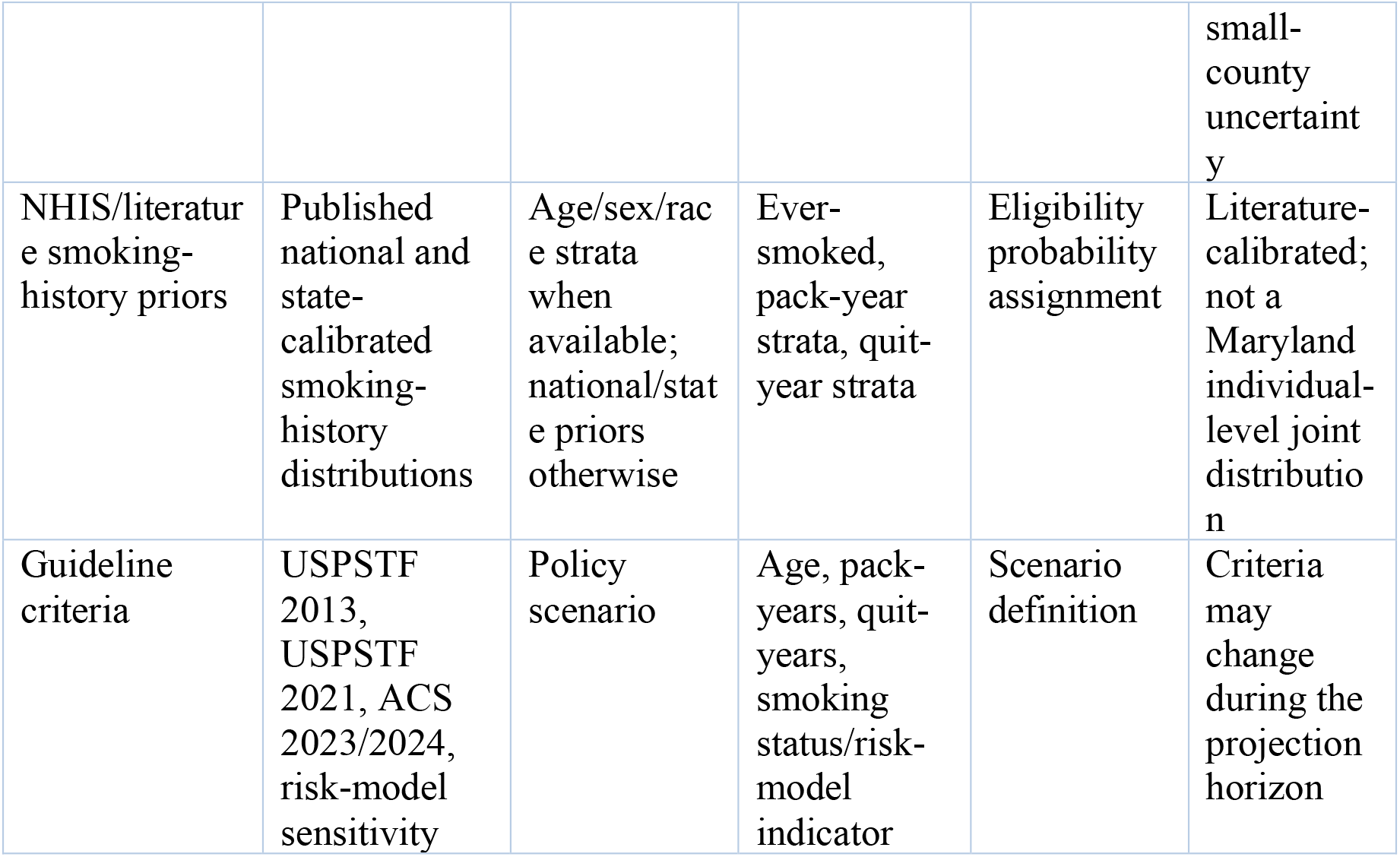
Source data and harmonization.

### Eligibility scenarios

We evaluated four screening criteria. They are the USPSTF 2013 legacy,^9^ USPSTF 2021 base case,^10^ ACS 2023/2024 expanded criteria,^21^ and a risk-model-expanded sensitivity criteria.^24^ USPSTF 2021 was the primary scenario and was defined as age 50–80 years, at least 20 pack-years, and current smoking or quitting within 15 years.^10^ ACS 2023/2024 retained age 50–80 years and at least 20 pack-years but removed the quit-year cutoff.^21^ The risk-model-expanded sensitivity is not a true PLCOm2012 implementation because individual-level variables needed to calculate PLCOm2012 risk scores were unavailable in the public projection spine.^24^

### Smoking-history calibration

Eligibility probabilities were assigned through smoking-history components rather than current smoking status alone.^18,19,25^ The model used priors for ever-smoked status, pack-year strata (<20, 20–29, ≥30), and quit-year strata (current, former quit ≤15 years, remote former quit >15 years). County current-smoking estimates informed a shrinkage-adjusted county offset, while age, sex, race/ethnicity, education, income, and nativity modifiers redistributed the statewide Maryland calibration across population cells.^26^ This design was chosen to reduce the between-county exaggeration that occurs when current smoking is used as a direct proxy for eligibility.

### Cumulative unique stock-flow framework

We describe a deterministic cohort-cell stock-flow simulation that tracks eligibility entry and exit across age, sex, race/ethnicity, county-equivalent jurisdiction, year, and screening scenario cells. It is CISNET-inspired in its dynamic smoking-history logic^25,27,28^ but is not a CISNET natural-history model and does not estimate cancer incidence, stage distribution, mortality benefit, false positives, overdiagnosis, screening uptake, or downstream diagnostic outcomes.

For each cell g, year t, and scenario s, E[g,t,s] denotes the prevalent eligible stock. The code computes E[g,t,s] = N[g,t] × p[eligible | g,t,s]. Let r[g,t,s] denote the annual exit fraction from mortality, age-out, and criterion-specific quit-window crossing, with r[g,t,s] = 1 − (1 − m[g,t]) × (1 − a[g,t,s]) × (1 − q[g,t,s]). The surviving prior-year eligible stock is S[g,t,s] = E[g,t−1,s] × (1 − r[g,t,s]). First-time eligible entrants are I[g,t,s] = max(0, E[g,t,s] − S[g,t,s]). Cumulative unique eligibility is initialized as U[g,2026,s] = E[g,2026,s] and updated as U[g,t,s] = U[g,t−1,s] + I[g,t,s] for t > 2026. State and county-equivalent totals were obtained by summing across cells.

### Subgroup analyses

Marginal subgroup counts were generated by age, sex, race/ethnicity, county-equivalent jurisdiction, education, household income, and nativity.^14,23^ These are marginal, not fully cross-classified, estimates, therefore, categories sum within each dimension but should not be summed across dimensions. Full subgroup tables are provided in the accompanying Data Supplement and package CSV files.

## Results

Because cumulative unique eligibility counts persons once at first eligibility, these values are not directly comparable with annual prevalent person-years. The latter are reported only to quantify the degree of overcounting.

Under the USPSTF 2021 base case,^10^ 238,346 Maryland residents were eligible for LDCT in 2026 and 245,326 in 2045. Cumulative unique eligibility over the period 2026 to 2045 was 768,668, compared with 4,850,735 annual prevalent person-years if annual counts were summed naively. The ACS 2023/2024 scenario^21^ increased the 2026 annual denominator to 314,616 and cumulative unique eligibility to 902,796 by adding remote former smokers with qualifying pack-year histories (Table 2, Figure 1).

**Table 2.** Statewide annual prevalent and 20-year cumulative unique eligibility by scenario.

| Scenario | 2026<br>annual | 2045<br>annual | Mean<br>annual | Cumulative<br>unique | Naive<br>annual<br>sum | Overcount<br>factor |
| --- | --- | --- | --- | --- | --- | --- |
| USPSTF<br>2013 legacy | 143,722 | 147,932 | 146,250 | 495,065 | 2,924,995 | 5.91x |
| USPSTF<br>2021 base<br>case | 238,346 | 245,326 | 242,537 | 768,668 | 4,850,735 | 6.31x |
| ACS<br>2023/2024<br>expanded | 314,616 | 323,830 | 320,148 | 902,796 | 6,402,970 | 7.09x |
| Risk-model-expanded<br>sensitivity | 226,428 | 233,060 | 230,410 | 747,782 | 4,608,198 | 6.16x |

**Figure 1.**
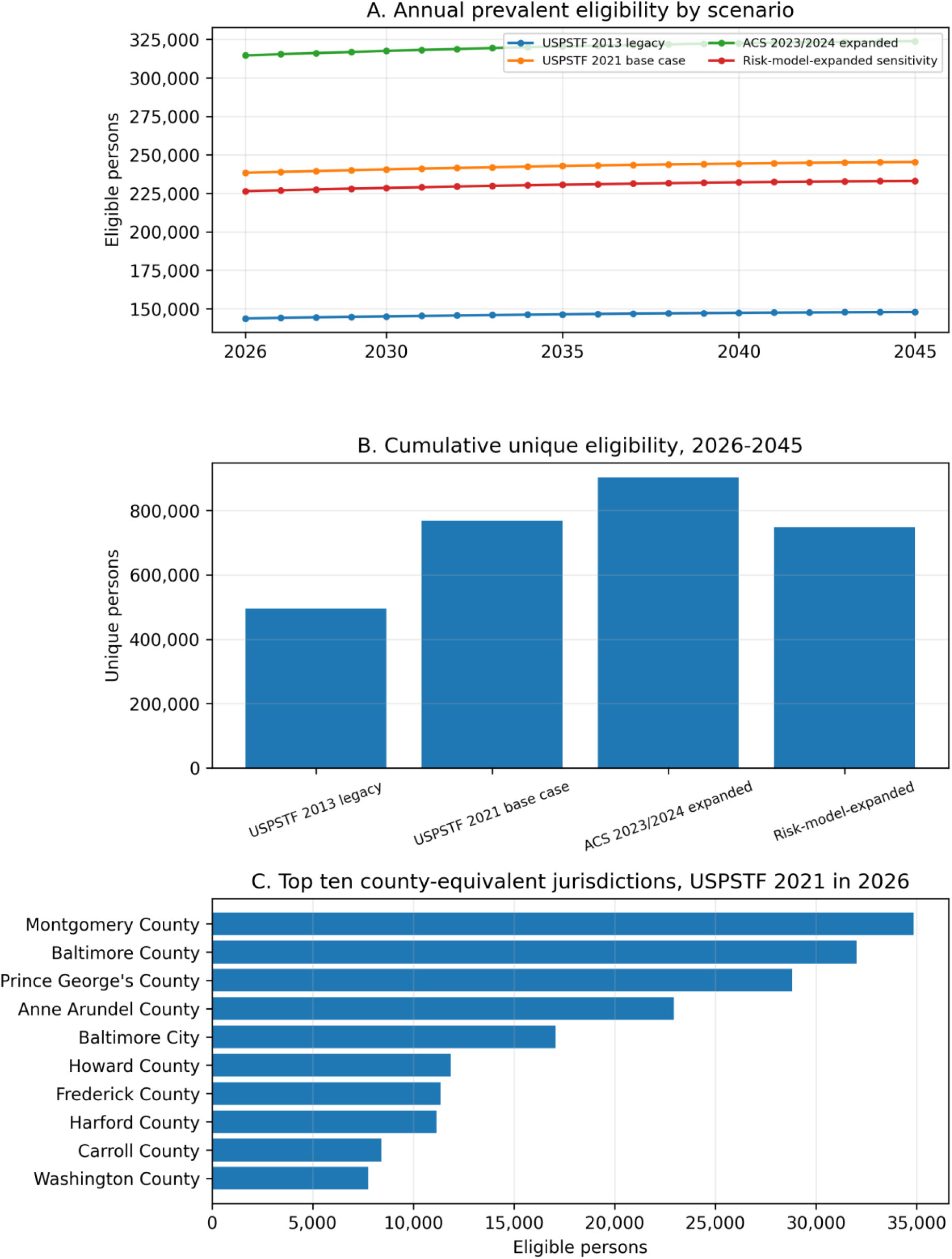
Planning outputs. A, Annual prevalent eligibility trajectories by screening scenario. B, Cumulative unique eligibility over 2026–2045. C, Top ten county-equivalent jurisdictions under USPSTF 2021 in 2026.

County-equivalent jurisdiction-level burden was concentrated in Maryland’s largest jurisdictions. Montgomery County, Baltimore County, Prince George’s County, Anne Arundel County, and Baltimore City contributed the largest 2026 annual eligible pools (Table 3). Complete jurisdiction-year estimates are provided in the Data Supplement and processed CSV files.

**Table 3.** Ten largest county-equivalent jurisdiction USPSTF 2021 annual eligibility pools in 2026.

| **Jurisdiction** | **2026**<br>**annual** | **2045**<br>**annual** | **Mean**<br>**annual** | **Cumulative**<br>**unique** |
| --- | --- | --- | --- | --- |
| Montgomery County | 34,840 | 35,902 | 35,473 | 112,426 |
| Baltimore County | 32,008 | 30,518 | 31,354 | 99,368 |
| Prince George's County | 28,803 | 30,513 | 29,744 | 94,267 |
| Anne Arundel County | 22,926 | 23,404 | 23,232 | 73,629 |
| Baltimore City | 17,055 | 15,604 | 16,377 | 51,903 |
| Howard County | 11,868 | 13,144 | 12,542 | 39,750 |
| Frederick County | 11,348 | 13,661 | 12,541 | 39,745 |
| Harford County | 11,147 | 11,594 | 11,404 | 36,141 |
| Carroll County | 8,416 | 8,348 | 8,406 | 26,642 |
| Washington County | 7,754 | 8,363 | 8,082 | 25,614 |

The ACS row is set-monotonic with USPSTF 2021. ACS current and recent-former components equal the USPSTF 2021 current and recent-former components, then add 76,270 remote former smokers (Table 4).

**Table 4.** Corrected 2026 pack-year and quit-year distribution for USPSTF 2021 and ACS 2023/2024.

| **Scenario** | **Smoking-**<br>**history**<br>**stratum** | **20-29 pack-**<br>**years** | **30-39**<br>**pack-years** | **>=40 pack-**<br>**years** | **Total** |
| --- | --- | --- | --- | --- | --- |
| **USPSTF**<br>**2021 base**<br>**case** | Current<br>smoker | 42,400 | 25,700 | 32,831 | 100,931 |
| **USPSTF**<br>**2021 base**<br>**case** | Former<br>smoker, quit<br><=15 y | 50,000 | 36,300 | 51,115 | 137,415 |
| **USPSTF**<br>**2021 base**<br>**case** | Former<br>smoker, quit<br>>15 y | 0 | 0 | 0 | 0 |
| **USPSTF**<br>**2021 base**<br>**case** | Total | 92,400 | 62,000 | 83,946 | 238,346 |
| **ACS**<br>**2023/2024**<br>**expanded** | Current<br>smoker | 42,400 | 25,700 | 32,831 | 100,931 |
| **ACS**<br>**2023/2024**<br>**expanded** | Former<br>smoker, quit<br><=15 y | 50,000 | 36,300 | 51,115 | 137,415 |
| **ACS**<br>**2023/2024**<br>**expanded** | Former<br>smoker, quit<br>>15 y | 29,000 | 19,000 | 28,270 | 76,270 |
| **ACS**<br>**2023/2024**<br>**expanded** | Total | 121,400 | 81,000 | 112,216 | 314,616 |

Subgroup and ever-smoked-only sensitivity results

Marginal subgroup estimates identified equity-relevant variation beyond age, sex, and race/ethnicity.^14^ Under USPSTF 2021 in 2026, estimated eligibility included 77,000 persons with high school/GED education, 70,000 with some college or associate degree, 74,000 with household income below $50,000, 82,000 with household income $50,000–$99,999, and 26,346 foreign-born residents; these values are marginal subgroup counts and should not be summed across dimensions.

The adult ever-smoked-only capacity stress test produced 1,957,699 eligible Maryland adults in 2026 and 3,383,683 cumulative unique persons over 2026–2045. Restricting ever-smoked-only eligibility to ages 50–80 produced 969,586 annual eligible residents in 2026 and 2,091,694 cumulative unique persons. These scenarios represent tobacco-exposed population denominators rather than recommended clinical LDCT eligibility criteria.

## Discussion

Our population-informatics model has significant implications for a statewide Maryland lung cancer screening initiative and other states that may pursue similar initiatives. First, the planning denominator depends strongly on whether the program needs annual prevalent capacity or cumulative unique reach. Under USPSTF 2021,^10^ the annual eligible population is approximately 238,000–245,000, but the 20-year cumulative unique population is approximately 769,000. Annual summation would overstate the unique-person denominator by more than sixfold. This distinction has direct consequences for programmatic capacity planning, workforce allocation, and cost forecasting.^27,29^

Second, explicit smoking-history modeling materially changes the planning estimate relative to current-smoking proxy analyses.^19,25,26^ Our framework includes former smokers with qualifying pack-year and quit-year histories and avoids allowing county current-smoking prevalence to dominate all eligibility variation. This shifts a larger share of burden toward populous and aging county-equivalent jurisdictions while preserving rural and high-smoking areas as priority equity geographies for access interventions.^16,17,20^

Third, eligibility policy changes can alter capacity requirements. ACS 2023/2024 criteria^21^ are a superset of USPSTF 2021 for adults aged 50–80 years with at least 20 pack-years because the quit-year exclusion is removed. Programs that build capacity only to USPSTF 2021 may underprepare for guideline convergence or payer expansion that includes remote former smokers.^30^ Risk-model-based approaches have been proposed to improve screening efficiency and reduce racial and ethnic disparities in eligibility;^13,24,31^ however, their population-level implementation requires individual-level risk factor data that are not available in public projection sources. Recent cost-effectiveness analyses support the value of expanding quit-year windows beyond the current 15-year cutoff.^29^

### Limitations

This analysis used public projection and surveillance inputs rather than individual-level smoking histories.^22,23^ Education, income, and nativity were represented as static marginal ACS covariates rather than annually projected joint distributions. The risk-model-expanded sensitivity is a planning sensitivity, not an individual PLCOm2012 score.^24^ The model estimates eligibility, not LDCT uptake, adherence, false positives, cancer incidence, stage shift, or mortality benefit.^5,7,27^ Finally, source data vintages and guideline criteria should be refreshed before operational launch and again when MDP projections or screening recommendations are updated.

## Data Availability

The submission package to the journal will include processed analytic CSV files, a source-data manifest, a Data Supplement, and reproducibility code in Python. External public data sources are documented with URLs, vintages, variables, roles in the model, and limitations. The package is designed to regenerate the submitted tables and figures from code-ready processed inputs.

## Data and code availability

The submission package includes processed analytic CSV files, a source-data manifest, a Data Supplement, and reproducibility code in Python. External public data sources are documented with URLs, vintages, variables, roles in the model, and limitations. The package is designed to regenerate the submitted tables and figures from code-ready processed inputs.

